# Long non-coding RNA *SNHG8* drives stress granule formation in tauopathies

**DOI:** 10.1101/2023.02.27.23286548

**Authors:** Reshma Bhagat, Miguel A. Minaya, Arun Renganathan, Muneshwar Mehra, Jacob Marsh, Rita Martinez, Alissa L. Nana, Salvatore Spina, William W. Seeley, Lea T. Grinberg, Celeste M. Karch

## Abstract

Tauopathies are a heterogenous group of neurodegenerative disorders characterized by tau aggregation in the brain. In a subset of tauopathies, rare mutations in the *MAPT* gene, which encodes the tau protein, are sufficient to cause disease; however, the events downstream of *MAPT* mutations are poorly understood. Here, we investigate the role of long non-coding RNAs (lncRNAs), transcripts >200 nucleotides with low/no coding potential that regulate transcription and translation, and their role in tauopathy. Using stem cell derived neurons from patients carrying a *MAPT* p.P301L, IVS10+16, or p.R406W mutation, and CRISPR-corrected isogenic controls, we identified transcriptomic changes that occur as a function of the *MAPT* mutant allele. We identified 15 lncRNAs that were commonly differentially expressed across the three *MAPT* mutations. The commonly differentially expressed lncRNAs interact with RNA-binding proteins that regulate stress granule formation. Among these lncRNAs, *SNHG8* was significantly reduced in a mouse model of tauopathy and in FTLD-tau, progressive supranuclear palsy, and Alzheimer’s disease brains. We show that *SNHG8* interacts with tau and stress granule-associated RNA-binding protein TIA1. Overexpression of mutant tau *in vitro* is sufficient to reduce *SNHG8* expression and induce stress granule formation. Rescuing *SNHG8* expression leads to reduced stress granule formation and reduced TIA1 levels, suggesting that dysregulation of this non-coding RNA is a causal factor driving stress granule formation via TIA1 in tauopathies.

## Introduction

Tauopathies are a class of neurodegenerative diseases that manifest as cognitive decline and are neuropathologically characterized by the accumulation of intracellular hyperphosphorylated tau protein (Bodea et al. 2016). Dominantly inherited mutations in the *MAPT* gene, which encodes the tau protein, are sufficient to cause disease in a subset of tauopathies termed frontotemporal lobar degeneration with tau pathology (FTLD-tau) (Pottier et al. 2016). However, the underlying mechanisms by which *MAPT* mutations cause disease remains unclear.

Several mechanisms contributing to FTLD-tau have been proposed. *MAPT* mutations have been reported to affect molecular and structural properties of tau. As a consequence, microtubule binding efficiency, post-translational modification status, and isoform balance of tau in the central nervous system (CNS) may be altered (Van Swieten and Spillantini 2007). *MAPT* mutations also lead to tau accumulation, impaired neuronal function, cell death, mitochondrial stress, autophagic and lysosomal dysregulation, and nuclear-cytosolic transport defects (Caballero et al. 2018; Frost, Bardai, and Feany 2016; Mahali et al. 2022; Pradeepkiran and Hemachandra Reddy 2020; Tracy et al. 2022; Zhu et al. 2017). Whether there are mechanisms upstream of these molecular events remains poorly understood.

Disruption of non-coding regulatory elements in the genome may have broad downstream effects that have yet to be fully explored in FTLD-tau (Simone et al. 2021; Yan et al. 2020). Stem cell modeling along with genome editing have revealed that *MAPT* mutations are sufficient to elicit a number of molecular events associated with synaptic function and proteostasis (Bowles et al. 2021; Hernandez et al. 2019; Jiang et al. 2018; Mahali et al. 2022; Minaya et al. 2023). These studies have focused on understanding the effects of *MAPT* mutations on coding genes. Yet, coding genes represent only 2% of the human genome. Non-coding regions, such as long non-coding RNAs (lncRNAs), represent 31.79% of the genome. LncRNAs play crucial regulatory roles in many cellular processes (Oo, Brandes, and Leisegang 2022), including the regulation of transcriptional modulation, post-transcriptional control, nuclear-cytoplasmic transport, translational inhibition, mRNA stability, RNA decoys, and regulation of protein activity (X. Zhang et al. 2019). LncRNAs also interact with a wide range of RNA-binding proteins, including those involved in stress granule formation (Khong et al. 2017; Van Treeck et al. 2018). With non-coding RNA making up a significant portion of the human genome, the impact of *MAPT* mutations on lncRNAs is an unexplored area that may hold key insights into the underlying mechanisms of FTLD-tau.

Our findings suggest that *MAPT* mutations have a significant impact on lncRNA expression in human neurons. We identified a lncRNA, *SNHG8*, that is reduced across three types of *MAPT* mutations and reduced in brains from tauopathy mouse models and human patients. *In vitro* studies demonstrate that *MAPT* mutations disrupt *SNGH8* expression, which promotes stress granule formation. This represents a novel mechanism that could be targeted for therapeutic intervention in the context of tauopathies. These results highlight the importance of studying the role of lncRNAs in the regulation of stress granule formation and the effects of *MAPT* mutations on lncRNA expression in the development of effective treatments for tauopathies.

## Material and Methods

### Patient consent

To obtain fibroblasts, skin punches were performed following written informed consent from the donor. The informed consent was approved by the Washington University School of Medicine Institutional Review Board and Ethics Committee (IRB 201104178 and 201306108). The University of California San Francisco Institutional Review Board approved the operating protocols of the UCSF Neurodegenerative Disease Brain Bank (from which brain tissues were obtained). Participants or their surrogates provided consent for autopsy, in keeping with the guidelines put forth in the Declaration of Helsinki, by signing the hospital’s autopsy form. If the participant had not provided future consent before death, the DPOA or next of kin provided it after death. All data were analyzed anonymously.

### iPSC generation and genome engineering

Human iPSCs used in this study have been previously described (Karch et al. 2019). iPSC lines were generated using non-integrating Sendai virus carrying the Yamanaka factors: OCT3/4, SOX2, KLF4, and cMYC (Life Technologies) (Ban et al. 2011; Takahashi and Yamanaka 2006). The following parameters were used for the characterization of each of the iPSC lines using standard methods (Takahashi and Yamanaka 2006): pluripotency markers by immunocytochemistry (ICC) and quantitative PCR (qPCR); spontaneous or TriDiff differentiation into the three germ layers by ICC and qPCR; assessment of chromosomal abnormalities by karyotyping; and *MAPT* mutation status confirmation by Sanger sequencing (characterization data previously reported (Minaya et al. 2023)).

To determine the impact of the *MAPT* mutant allele on molecular phenotypes, we used CRISPR/Cas9-edited isogenic controls in which the mutant allele was reverted to the wild-type (WT) allele in each of the donor iPSC lines as previously described (Karch et al. 2019; Minaya et al. 2023). The resulting edited iPSC lines were characterized as described above in addition to on- and off-target sequencing (characterization data previously reported (Minaya et al. 2023)). All iPSC lines used in this study carry the *MAPT* H1/H1 common haplotype. All cell lines were confirmed to be free of mycoplasma.

### Differentiation of iPSCs into cortical neurons

iPSCs were differentiated into cortical neurons as previously described (Karch et al. 2019; Mahali et al. 2022)(https://dx.doi.org/10.17504/protocols.io.p9kdr4w). Briefly, iPSCs were plated at a density of 65,000 cells per well in neural induction media (StemCell Technologies) in a 96-well v-bottom plate to form neural aggregates. After 5 days, cells were transferred into culture plates. The resulting neural rosettes were isolated by enzymatic selection (Neural Rosette Selection Reagent; StemCell Technologies) and cultured as neural progenitor cells (NPCs). NPCs were differentiated in planar culture in neuronal maturation medium (neurobasal medium supplemented with B27, GDNF, BDNF, and cAMP). The cells were analyzed after 6 weeks in neuronal maturation medium. At this time, tau protein levels are stable and similar to protein profiles described in human brains (Sato et al. 2018).

### RNA sequencing and lncRNA transcript quantification

RNAseq was generated from iPSC-derived neurons as previously described (Jiang et al. 2018; Minaya et al. 2023). Briefly, samples were sequenced by an Illumina HiSeq 4000 Systems Technology with a read length of 1×150 bp and an average library size of 36.5 ± 12.2 million reads per sample.

We used Salmon (v. 0.11.3) (Patro et al. 2017) to quantify the expression of the genes annotated within the human reference genome (GRCh38.p13; **Supplemental Table 1**). The lncRNA genes were selected for downstream analyses. LncRNA genes that were present in at least 10% of samples with expression >0.1 TPM were included in subsequent analyses: 7,537 lncRNA genes (**Supplemental Figure 1**).

### Principal component and differential expression analyses

Principal component analyses (PCA) were performed with the selected 7,537 non-coding genes using regularized-logarithm transformation (rlog) counts. Differential gene expression was performed using the DESeq2 (v.1.22.2) R package (Love, Huber, and Anders 2014). PCA and differential gene expression analyses were performed independently for each pair of *MAPT* mutations and isogenic controls. Each *MAPT* mutation and its isogenic control were considered independent cohorts due to their shared genetic background. PCA and Volcano plots were created for each comparison using the ggplot2 R package (v3.3.6) (Wilkinson 2011).

### Functional annotation of differentially expressed lncRNA genes

LncSEA was used to determine the RNA-binding protein interactions of common differentially expressed lncRNAs (Chen et al. 2021). Gene relationships of top RNA-binding proteins and *MAPT*, including physical interaction, co-localization, pathway, shared protein domain, and genetic interaction, were examined using the GeneMANIA Cytoscape plugin (Montojo et al. 2010).

CatRAPID was applied to identify the interactions between individual lncRNAs and RNA-binding proteins (Armaos et al. 2021; Bellucci et al. 2011). The input for CatRAPID analysis was the FASTA sequence of lncRNA and protein. The output was a heat map where the axes represent the indexes of the RNA and protein sequences with interaction propensity and discriminative power. The Interaction Propensity is a measure of the interaction probability between one protein (or region) and one RNA (or region). This measure is based on the observed tendency of the components of ribonucleoprotein complexes to exhibit specific properties of their physio-chemical profiles that can be used to make a prediction. The Discriminative Power is a statistical measure introduced to evaluate the Interaction Propensity with respect to CatRAPID training. It represents confidence of the prediction. The Discriminative Power (DP) ranges from 0% (unpredictable) to 100% (predictable). DP values above 50% indicate that the interaction is likely to take place, whereas DPs above 75% represent high-confidence predictions.

### Plasmids

Plasmids pRK5-EGFP containing 4R0N Tau WT or P301L (Addgene plasmids 46904 and 46908) were used to evaluate the impact of tau on stress granule formation and lncRNA expression (Hoover et al. 2010). To test the impact of *SNHG8* on stress granule formation, a plasmid containing human *SNHG8* (transcript 203) in pcDNA3.1(+)-C-eGFP was used (pcDNA3.1(+)-SNHG8-203-EGFP (transcript 203) and control pcDNA3.1(+)-EGFP; Genescript). Untagged P301L-Tau (4R2N) constructs in pcDNA3.1(+) were employed in SNHG8-EGFP rescue experiments (Karch, Jeng, and Goate 2012).

### Transient Transfection of HEK293-T cells

HEK293-T cells were grown in Dulbecco’s Modified Eagle Medium (DMEM) (Life Technologies) supplemented with 10% FBS, 1% L-Glutamine, and 1% Penicillin and streptomycin solution. Plasmids were transfected using Lipofectamine 2000 (Invitrogen, San Diego, CA, USA) according to the manufacturer’s protocol. The transfected cells were evaluated after 24 or 48 hours for immunocytochemistry or RNA-immunoprecipitation, respectively.

### RNA Immunoprecipitation

To measure the binding between tau and *SNHG8*, RNA immunoprecipitation was performed as previously described with minor modifications (Abudayyeh et al. 2017; Kaewsapsak et al. 2017). Briefly, HEK293-T cells were grown in a T175 flask at a density of approximately 80%, and cells were transfected with pRK5-EGFP-4R0N-Tau WT (10ug) or pRK5-EGFP control (10ug). After 48 hours, cells were fixed with 0.1% paraformaldehyde in PBS for 10 minutes at room temperature and then washed with 125 mM glycine in PBS for 5 minutes. Cells were lysed in RIPA buffer (50mM Tris-HCl pH 7.4, 100mM NaCl, 1% Igepal CA-630, 0.1% SDS, 0.5% Sodium Deoxycholate) supplemented with protease inhibitor mixture (Sigma-Aldrich) and RNase inhibitor before centrifugation at 16,000×*g* for 10 minutes at 4°C. 5% of the supernatant was saved as input, and the remaining clarified lysate was then incubated with 40μL anti-GFP magnetic beads (ChromoTek GFP-Trap Agarose Nanobody (VHH), gtmak-20) overnight at 4°C with rotation. The beads were then pelleted and washed four times with high salt buffer (50mM Tris-HCl pH-7.4, 1M NaCl, 1mM EDTA, 1% Tgepal CA-630, 0.1% SDS and 0.5% sodium deoxycholate) supplemented with 0.02% Tween-20. Enriched RNAs were released from the beads in 200μL of Trizol. Thereafter, RNA was isolated from the IP-eluted RNA and from 5ul input samples using standard RNeasy Mini Kit (250) (Qiagen-74106) following the manufacturer’s protocol. The resulting IP-eluted RNA and input RNA is used as the template for cDNA synthesis using SuperScript III (Invitrogen) and measured by qPCR on a Quantstudio 3 qPCR machine (Applied Biosystems by Thermo Fisher Scientific) using specific primers for *SNHG8*. The recovery of specific RNAs was calculated by dividing transcript abundance in the enriched sample by the corresponding input.

### Mouse model of tauopathy

To evaluate whether the genes differentially expressed in iPSC-derived neurons from *MAPT* mutation carriers were altered in animal models of tauopathy, we analyzed transcriptomic data from a Tau-P301L mouse model of tauopathy and non-transgenic controls (Matarin et al. 2015; Ramsden et al. 2005). Differential gene expression of lncRNAs was performed in mice at 2, 4, and 8 months of age using unpaired t-tests to assess significance.

### Gene expression analysis in PSP and AD brains

To determine whether the differentially expressed lncRNAs in the *MAPT* mutant iPSC-derived neurons capture molecular processes that occur in human brains with primary tauopathy, we analyzed gene expression in a publicly available dataset: the temporal cortex of 76 control, 82 PSP, and 84 AD brains (syn6090813) (Allen et al. 2016). Differential gene expression analyses comparing controls with PSP and AD brains were performed using a “Simple Model” that employs multi-variable linear regression analyses using normalized gene expression measures and corrected by sex, age-at-death, RNA integrity number (RIN), brain tissue source, and flowcell as covariates (Allen et al. 2016). Transcriptomic data from the middle temporal gyrus of FTLD-tau patients with *MAPT* IVS10+16 and p.P301L mutation (*MAPT* IVS10+16 n=2 and *MAPT* p.P301L n=1) and neuropathology free controls (n=3) were also analyzed (Minaya et al. 2023). Differential expression analyses comparing FTLD-tau mutation carrier brains with controls was performed using DESeq2 (v.1.22.2) R package (Love, Huber, and Anders 2014) as previously described (Minaya et al. 2023).

### Induction and quantification of stress granules

TIA1 was used to monitor stress granule formation (Gilks et al. 2004). Stress granule formation was induced by culturing HEK293-T cells in a nutrient poor buffer (Hank’s buffer) as previously described (Hanson and Mair 2014; Kedersha and Anderson 2007). HEK293-T or iPSC-derived neurons were immunostained with a TIA1 (Sigma Aldrich-SAB4301803, 1:250 dilution) or G3BP2 (Cell Signaling Technology-31799S, 1:500 dilution) antibodies. Briefly, to perform immunocytochemistry, cells were grown on chamber slides. Culture media was aspirated, and cells were washed with PBS and fixed with 4% paraformaldehyde (Sigma) for 20 minutes at room temperature. Cells were washed with PBS and incubated with permeabilization buffer (0.1% Triton X-100 in PBS). Cells were then blocked in 0.1% bovine serum albumin (BSA; Sigma) and treated with primary and secondary antibodies diluted in 0.1% BSA. Immunostained cells were imaged (BZ-X800 series, Keyence fluorescent microscope, Keyence, IL, USA). At least six random images were captured per replicate, per condition. To calculate the percentage of cells positive for stress granules, the number of cells with stress granules in GFP-positive cells were divided by total number of GFP-positive cells. To determine the number of stress granules/cell and total stress granules in all GFP-positive cells, TIA1-positive inclusions were manually counted and corrected for the GFP-positive cells.

### RNAscope

RNAscope (Advanced Cell Diagnostics, ACD; Hayward, CA) was performed using BaseScope Reagent Kit v2 – RED (323900) kit by using specific probes targeting human *SNHG8* (NC_000004.12) according to the manufacturer’s protocol. 3ZZ probe named BA-Hs-SNHG8-O1-3zz-st targeting 2-133 of NC_000004.12:118278708-118279137 was used. BaseScope is a chromogenic assay: red chromogen was used for *SNHG8* detection which can be seen under a fluorescent microscope in the ∼Texas Red spectrum. HEK293-T cells were fixed with formaldehyde and washed with PBS. Slides were then hybridized with target probes and incubated in a HybEZ oven (ACD) for 2 hours at 40°C. Next, signals were amplified and generated with a BaseScope Detection Reagent Kit v2 – RED. Cells were then counterstained with DAPI. *SNHG8* expression was scored as positive if staining was present in HEK293-T cells. For visualizing the slides stained with *SNHG8*, a Keyence microscope (BZ-X800 series, Keyence fluorescent microscope, Keyence, IL, USA) was used. Images were captured at 40X magnification. ImageJ (https://imagej.nih.gov/) was used to quantify the mean intensity of *SNHG8*. The freehand selections tool was used to mark the transfected cells in green channel and the measure tool was used to quantify the *SNHG8* signal in the red channel.

### Statistical Analysis

Statistical analyses of biochemical and immunocytochemistry experiments were performed using GraphPad Prism version 9.2.0 (332) software. Each experiment was performed at least three times to determine statistical significance. Data distribution was assumed to be normal. Comparison between experimental and control group was analyzed using Student’s t-test, a level of p< 0.05 was considered statistically significant. Details of the sample sizes and statistical tests used are indicated in the figure legends.

## Results

### MAPT mutations are sufficient to drive changes in lncRNAs in human neurons

To explore the contribution of lncRNAs to tauopathy, we examined whether there were a common set of lncRNAs that are downstream of *MAPT* mutations. The more than 50 *MAPT* mutations fall into three major classes: (1) intronic mutations that alter splicing, leading to an imbalance in tau isoforms; (2) missense mutations within exon 10, leading to mutations in only a subset of tau isoforms; (3) missense mutations occurring in all tau isoforms. To begin to define the common non-coding mechanisms driving FTLD-tau, we have studied *MAPT* mutations that fall into each of the three major classes: *MAPT* IVS10+16, p.P301L, and p.R406W, respectively. Transcriptomic data from iPSC–derived neurons carrying *MAPT* IVS10+16, p.P301L, or p.R406W together with their CRISPR/Cas9-generated isogenic controls were analyzed (**Figure 1A**).

**Figure 1:**
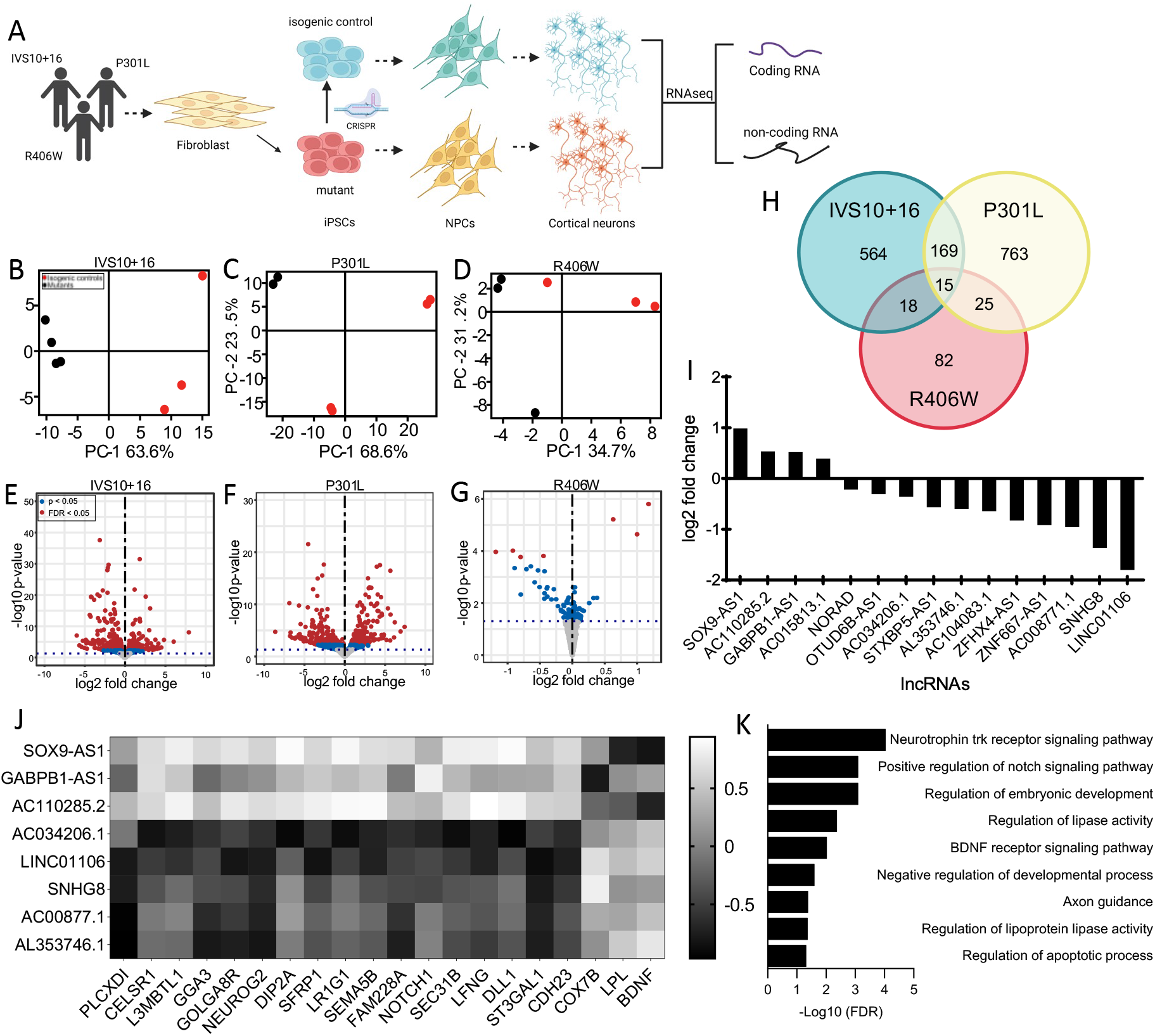
Mutations in *MAPT* are sufficient to drive changes in long non-coding RNA profile in iPSC-derived neurons. A. Diagram of experimental design. B-D. Principal component analysis (PCA) of *MAPT* IVS10+16, p.P301L, and p.R406W carriers and their respective isogenic controls using only long non-coding RNAs. Red dots, CRISPR-corrected isogenic controls. Black dots, *MAPT* mutation carriers. E-G. Volcano plots representing the differential expression of long non-coding RNAs in *MAPT* IVS10+16, p.P301L, and p.R406W carriers compared to their respective isogenic controls. Red dots, differentially expressed genes (FDR<0.05). Blue dots, differentially expressed genes (p<0.05). Grey dots, not significant. H. Venn diagram showing lncRNA overlap among all three *MAPT* mutations. I. Bar graph representing mean log2 foldchange of common differentially expressed lncRNAs. J. Heat map of correlation between differentially expressed lncRNAs and differentially expressed protein coding RNA. Correlation coefficient >0.6. K. GO terms from the analysis of highly correlated protein coding RNAs.

To define the global impact of *MAPT* mutations on lncRNAs across the genome, we performed differential expression analyses. Principal component analysis revealed that variation in the lncRNA transcriptome was sufficient to distinguish *MAPT* mutations from their isogenic controls (**Figure 1B-D**). Differential expression analyses identified a number of lncRNAs changing as a function of the presence of the mutant allele (**Figure 1E-G**; **Supplemental Table 2**; p<0.05, *MAPT* IVS10+16 (n=766; **Supplemental Table 3**), p.P301L (n=972; **Supplemental Table 4**), and p.R406W (n=141; **Supplemental Table 5**)). Among these, 15 lncRNAs were significantly altered in the same direction across the three datasets (**Figure 1H-I; Supplemental Table 6**). Thus, *MAPT* IVS10+16, p.P301L, and p.R406W mutations were sufficient to shift the lncRNA transcriptomic state of human neurons, and three classes of *MAPT* mutations generated a common molecular signature that we sought to further explore for their role in pathologic processes.

### Mutant tau-regulated lncRNAs disrupt the expression of coding genes in human neurons

To begin to define the regulatory role of the 15 common lncRNAs, we evaluated the impact of these lncRNAs on coding gene expression. We have previously reported that these three classes of *MAPT* mutations are sufficient to drive altered gene expression of 275 protein coding genes (p<0.05; (Minaya et al. 2023). LncRNAs can act in a cis or trans manner to inhibit or activate transcription of protein coding genes (Statello et al. 2021). We defined the coding genes that are proximal (<5 kb) to the lncRNAs (**Supplemental Table 7**). We also asked whether expression of the 15 common lncRNAs were correlated with expression of the 275 protein coding genes (**Figure 1J)**. We found that 8 of the 15 lncRNAs were highly correlated with 20 of the 275 protein coding genes: *SOX9-AS1, GABPB1-AS1, AC110285*.*2, AC034206*.*1, LINC01106, SNHG8, AC00877*.*1, AL353746*.*1* (**Figure 1J; Supplemental Table 8**). Gene enrichment analyses were then performed to determine the biological role of these regulatory relationships. The protein coding genes highly correlated with the differentially expressed lncRNAs were found to be enriched in pathways related to Neurotrophin trk receptor signaling (FDR = 9.22 × 10^−5^), Notch signaling (FDR = 4.8 × 10^−2^), BDNF signaling (FDR = 9.5 × 10^−3^), lipoprotein lipase activity (FDR = 4.3 × 10^−3^), and axonal guidance (FDR = 4.17 × 10^−2^) (**Figure 1K**). Together, these findings suggest that lncRNAs commonly altered by *MAPT* mutations exhibit broad gene regulatory roles.

### Mutant tau-regulated lncRNAs are enriched in RNA-binding proteins that function in stress granule formation

LncRNAs can also act as scaffolds or decoys to promote or weaken the interaction between macromolecules (K. C. Wang and Chang 2011). The interaction of lncRNAs with RNA-binding proteins affects posttranslational modifications, stability, subcellular localization, and activity of interacting partners (Yang, Wen, and Zhu 2015). RNA-binding proteins typically consist of aggregation-promoting, low complexity domains, or prion-like domains, and are involved in the formation of stress granules (Ash et al. 2021; Gerstberger et al. 2014; Latimer et al. 2021; Montalbano et al. 2020; Urwin et al. 2010). Tau interacts with a number of RNA-binding proteins *in vitro* and *in vivo* that then facilitate stress granule formation, which may act as precursors to tau aggregates in FTLD-tau (Lester et al. 2021).

To further examine the molecular functions of the 15 common lncRNAs, we evaluated the potential interaction with RNA-binding proteins using the lncSEA algorithm (Chen et al. 2021). The mutant tau-regulated lncRNAs were found to interact with 255 RNA-binding proteins, the top 15 are shown in **Figure 2A** (see also **Supplemental Table 9**). Interestingly, FUS, DDX3X, TARDBP (encoding TDP-43 protein), and TIA1 were predicted to be the most significant interacting partners (**Figure 2A**). *FUS* and *TARDBP* are FTLD genes, and DDX3X and TIA1 have been implicated in tauopathies (Ash et al. 2021; Lennox et al. 2020; Montalbano et al. 2020; Urwin et al. 2010). Gene network analysis of *FUS, DDX3X, TARDBP, TIA1*, and *MAPT Z*eveal that *TIA1, FUS, YBX1, ATXN2, APOE, MAP2, PWP2, APAF1, HNRNPA2B1*, and *ILF3* physically interact with *MAPT* (**Figure 2B**). Interactions between *MAP4, DDX3X, FUS, TARDBP, HNRNPA2B1, ILF3*, and *MAPT* have been experimentally validated (**Figure 2B**). *FUS, DDX3X, TARDBP, TIA1*, and *MAPT* function in pathways related to the regulation of response to stress (FDR = 1 × 10^−4^), regulation of translation (FDR = 1.3 × 10^−4^), regulation of autophagy (FDR = 1 × 10^−3^), and stress granule assembly (FDR= 1.06 × 10^−2^; **Figure 2C**). Thus, the 15 common lncRNAs interact with RNA-binding proteins that mediate stress granule formation.

**Figure 2:**
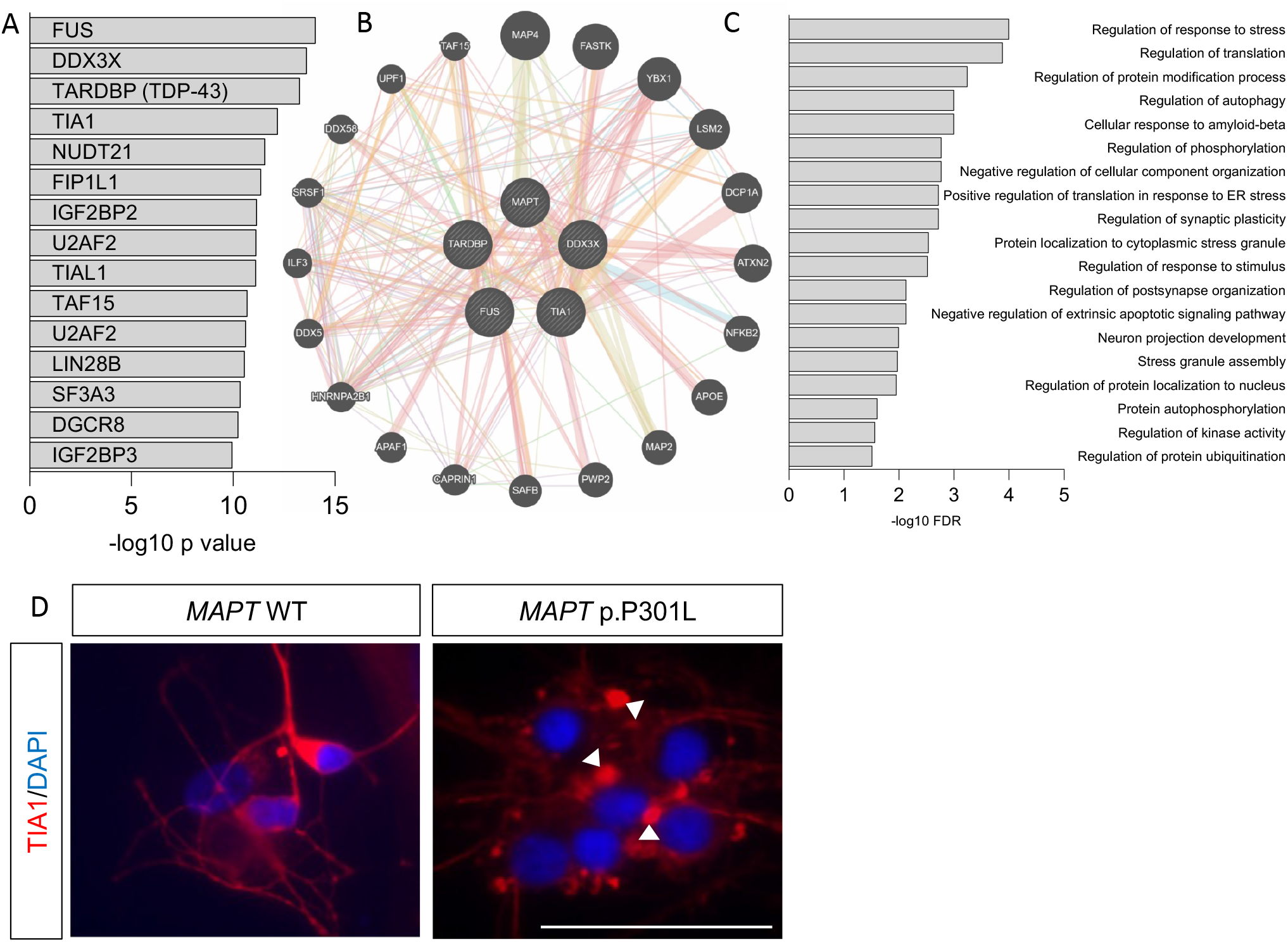
Common differentially expressed lncRNAs interact with RNA-binding proteins and are involved in stress granule formation. A. Common differentially expressed lncRNAs are predicted to interact with RNA-binding proteins interacting. B. GeneMANIA network of *FUS, DDX3X, TARDBP*, TIA1, and *MAPT*. C. GO terms obtained from the network using STRING. D. TIA1-positive stress granules are detectable in *MAPT* p.P301L neurons. White arrows indicate stress granules. Scale bar, 50µm.

Given our observation that common differentially expressed lncRNAs are enriched in RNA-binding proteins that regulate stress granule formation, we evaluated stress granule formation in iPSC-derived neurons from *MAPT* p.P301L and isogenic controls using immunocytochemistry for stress granule marker, TIA1 (**Figure 2D**). We observed *MAPT* mutant neurons have a marked increase in TIA1-positive stress granules. This is consistent with prior reports in mouse models of tauopathy (P301L-Tg4510 and PS19) and human FTLD-tau patients (B. F. Maziuk et al. 2018; Vanderweyde et al. 2016). Thus, *MAPT* mutations lead to the accumulation of stress granules in human neurons.

### SNHG8 is dysregulated in a mouse model of tauopathy and in human brain tissue

Despite the many strengths of human stem cell models, there remains a need to validate key discoveries in a dish using *in vivo* models to focus on those changes that are relevant to disease phenotypes. Thus, we sought to determine the extent to which the 15 commonly differentially expressed lncRNAs across *MAPT* mutations (**Figure 1H-I**) were altered as tau accumulates in the Tau-P301L mouse model of tauopathy. Using the Mouse Dementia Network (Matarin et al. 2015), we analyzed transcriptomic data generated from the cortex of WT and Tau-P301L mice collected at 2, 4, and 8 months (Ramsden et al. 2005)(**Figure 3A**). Among the 15 lncRNAs, only *Norad* (2900097C17Rik) and *Snhg8* were present in the dataset (**Supplemental Table 10**). The remaining 13 lncRNAs exhibit lower conservation among mammals and were not identified in the dataset. *Norad* expression was similar between WT and Tau-P301L at 2 and 4 months but was significantly elevated in Tau-P301L brains at 8 months, discordant with the findings in iPSC-neurons (**Supplemental Figure 2**). At 2 months of age, *Snhg8* expression was similar between WT and Tau-P301L (**Figure 3A**), suggesting that altered *Snhg8* expression was not developmentally encoded. At 4 and 8 months, *Snhg8* was significantly reduced in Tau-P301L mice (**Figure 3A**). The reduced expression of *Snhg8* at 4 and 8 months of age coincides with periods of active accumulation of tau aggregation in Tau-P301L mice (Matarin et al. 2015).

**Figure 3:**
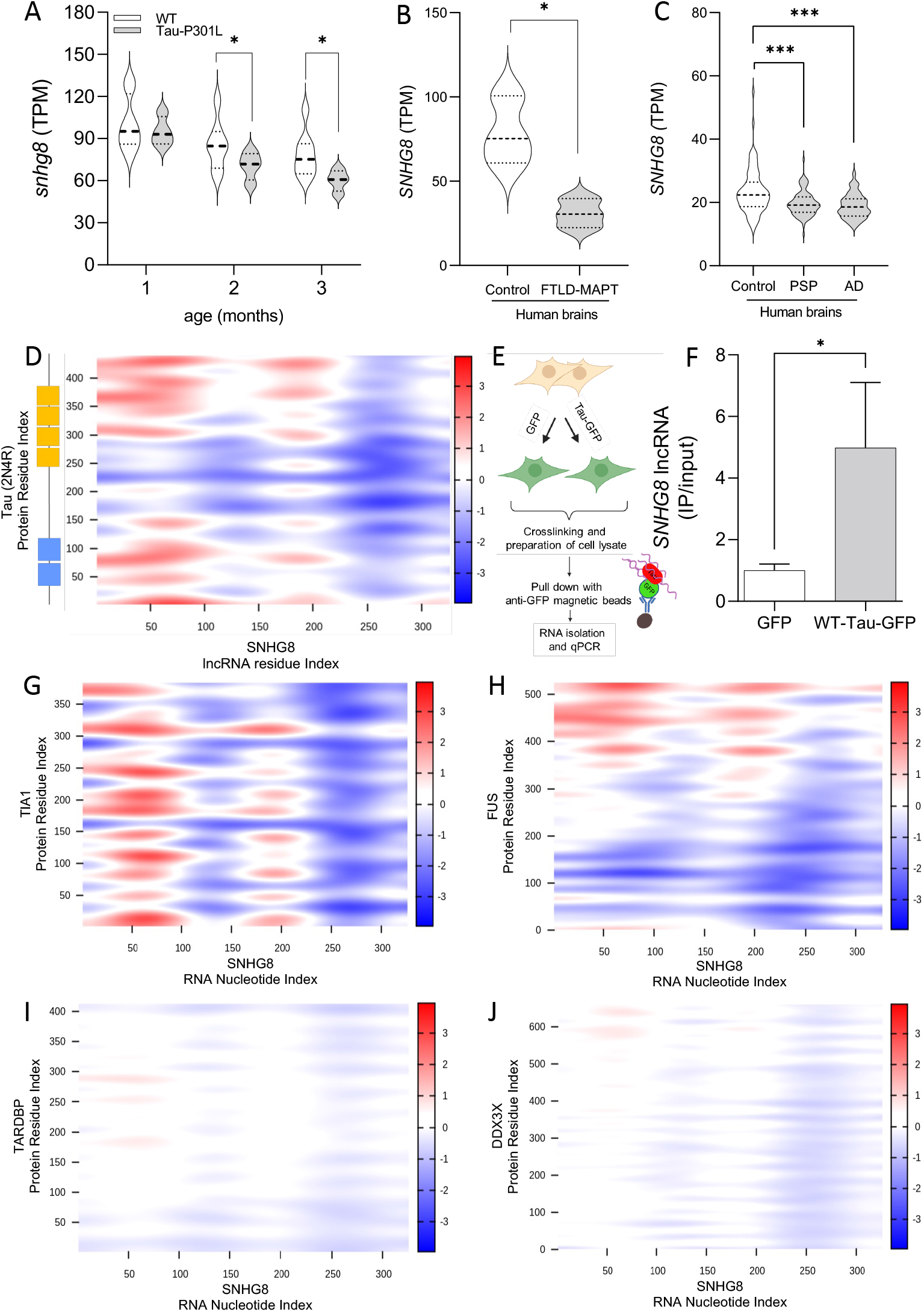
*SNHG8* is downregulated in mouse and human brains and interacts with tau, TIA1, FUS, TARDBP, and DDX3X. A. Normalized read counts (TPM) of *Snhg8* in WT mice and Tau-P301L mice. B. Normalized read counts of *SNHG8* in control and FTLD-tau (FTLD-MAPT, *MAPT* IVS10+16 and *MAPT* p.P301L carrier) brains. C. Normalized read counts of *SNHG8* in control, progressive supranuclear palsy (PSP), and Alzheimer’s disease (AD) brains. *, p≤0.05. ***, p≤0.001. D. CatRAPID interaction profile of 2N4R tau reveals multiple interaction domains with *SNHG8*. E. Schematic of the RNA pull down to measure the interaction between tau and *SNHG8*. F. Plot showing relative transcript expression from GFP pull down of GFP and WT-Tau-GFP transfected HEK293-T cells. Data are representative of 3 independent experiments. Bar graph is represented as mean ± SEM. Student’s paired t-test was used to determine significance. *, p≤0.05. G-J. CatRAPID interaction profile of tau or RNA-binding proteins with *SNHG8*, the colors in the heatmap indicate the interaction score (ranging from -3 to +3) of the individual amino acid and nucleotide pairs.

Leveraging isogenic iPSC lines to understand the contribution of a single allele to downstream phenotypes is a powerful system that, when applied here, has revealed lncRNAs shared across *MAPT* mutations that also change with tau accumulation in mouse models of tauopathy. However, a limitation of this approach is that iPSC-neurons are cultured in a dish and remain relatively immature. For example, iPSC-neurons predominantly express 0N3R tau (Patani et al. 2012; Sato et al. 2018; Sposito et al. 2015), while the adult brain expresses 6 tau (Hefti et al. 2018; Sato et al. 2018). Thus, we sought to determine whether *SNHG8* is altered in human brains of tauopathy patients. *SNHG8* was significantly reduced in FTLD-tau caused by *MAPT* IVS10+16 or MAPT p.P301L (**Figure 3B, Supplemental Table 11**); a primary sporadic tauopathy progressive supranuclear palsy (**Figure 3C, Supplemental Table 12**); and a secondary tauopathy Alzheimer’s disease (**Figure 3C, Supplemental Table 13**). Together, we show *SNHG8* is altered in mouse models of tauopathy and in tauopathy patient brains, supporting a role for *SNHG8* in pathologic processes.

### SNHG8 interacts with tau and RNA-binding proteins

Given our findings of *SNHG8* dysregulation in iPSC-derived neurons from *MAPT* mutation carriers, mouse brains with tau aggregation, and human brains from tauopathy patients, we asked whether this effect occurs via direct interaction between tau and *SNHG8*. CatRAPID, a bioinformatic platform that predicts interactions between protein and RNA based on structure data was employed (Bellucci et al. 2011). The longest tau isoform (2N4R) is predicted to have several binding domains with *SNHG8* (**Figure 3D**). To functionally validate the predicted interaction between tau and *SNHG8 in vitro*, HEK293-T cells were transiently transfected with plasmids containing WT-Tau-GFP or a GFP control, cells were crosslinked to retain binding partners, and tau was enriched by immunoprecipitation for GFP (**Figure 3E**). qPCR of the RNA fraction bound to GFP revealed a 5-fold enrichment of *SNHG8* in the WT-Tau-GFP fraction compared with the GFP control fraction (p<0.05; **Figure 3F**). Thus, *SNHG8* interacts with tau protein.

Common differentially expressed lncRNAs were enriched in RNA-binding proteins TIA1, FUS, DDX3X, and TDP-43 (**Figure 2A**); so, we asked whether *SNHG8* interacts with these RNA-binding proteins and where the interaction occurs. *SNHG8* is predicted to bind to regions of TIA1, FUS, DDX3X, and TDP-43 (**Figure 3G-J**). TIA1 showed the strongest interaction with *SNHG8* (interaction propensity: 65; **Figure 3G**). FUS, DDX3X, and TDP-43 with *SNHG8* to a lesser extent but passed the threshold for positive interaction (interaction propensity: 45, 37, and 14, respectively; **Figure 3H-J**).

### Mutant tau and stress drive stress granule formation via SNHG8

TIA1 has been shown to interact with tau and to facilitate incorporation into stress granules (Piatnitskaia et al. 2019; Vanderweyde et al. 2016). To evaluate the impact of a representative *MAPT* mutation on stress granule formation *in vitro*, we compared HEK293-T cells in which plasmids containing WT-Tau-GFP or P301L-Tau-GFP were transiently overexpressed (**Figure 4A**). TIA1, a stress granule resident protein, was used as a marker of stress granule accumulation. Under basal conditions, the percentage of tau-positive cells with TIA1-positive stress granules and the number of stress granules per cell was significantly enriched in the P301L-Tau-GFP expressing cells compared with WT-Tau-GFP (p<0.05; **Figure 4B-D**).

**Figure 4:**
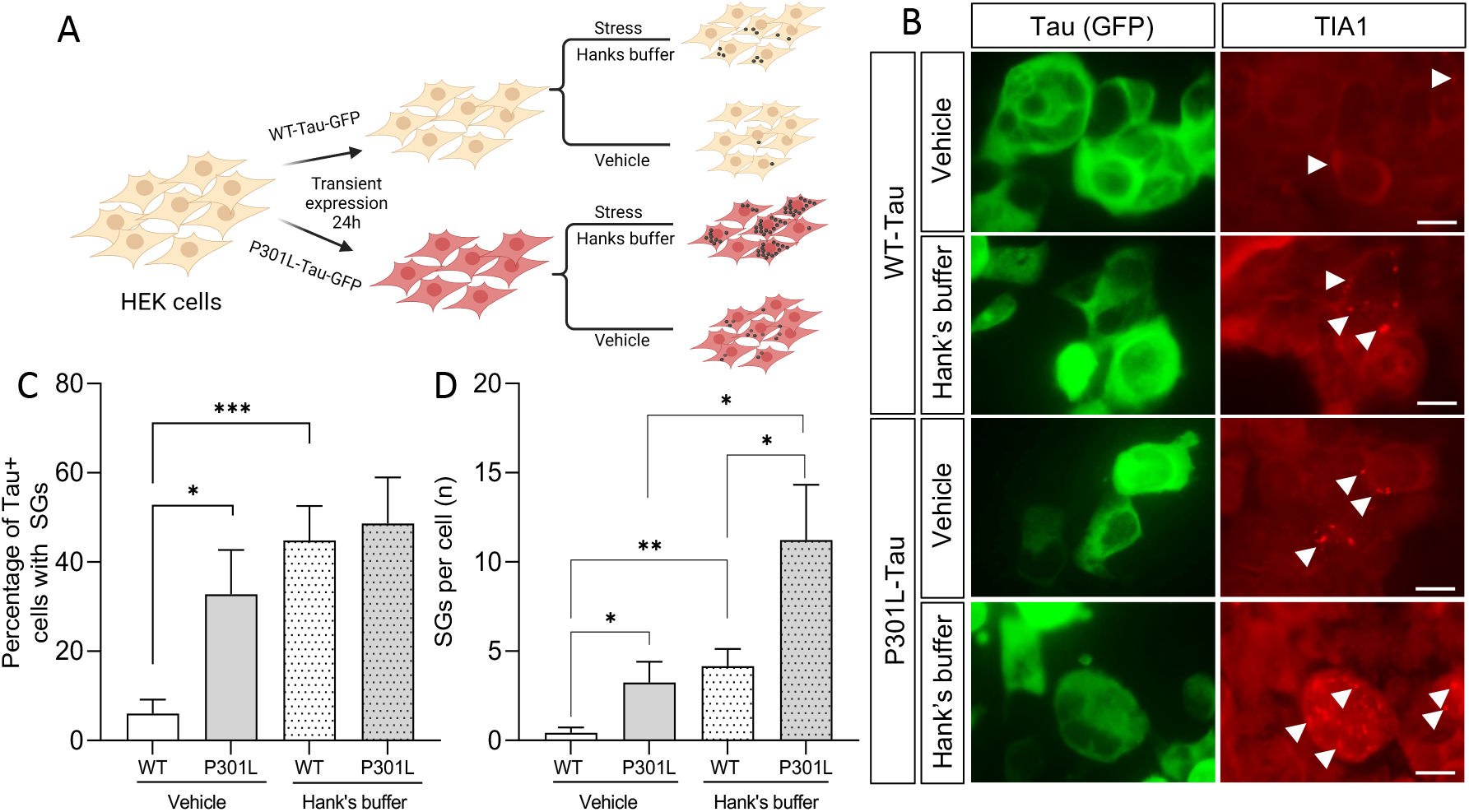
Mutant tau and starvation induce stress granule formation. A. Schematic of paradigm. B. Immunocytochemistry of HEK293-T cells with TIA1 antibody under basal (vehicle) and stress (Hank’s buffer) conditions. GFP (green), TIA1 (red). White arrow, stress granules. Scale bar, 50um. C. Graph representing the quantification of cells with TIA1-positive stress granules. D. Graph of the number of stress granules in GFP-positive cells. The data represents 4 independent experiments. Bar graphs represent mean ± SEM. Statistical significance determine with a Student’s t-test. *, p≤0.05; **, p≤0.001, and ***, p≤0.0001.

Next, we explored whether there was a differential genotypic impact on stress granule formation with stress introduced by starvation (**Figure 4A**). HEK293-T cells were transfected with WT-Tau-GFP or P301L-Tau-GFP and cultured in Hank’s buffer for 1 hour prior to immunocytochemistry. Stress induction by starvation was sufficient to produce an increase in the percentage of tau-positive and TIA1-positive stress granules in WT-Tau-GFP and P301L-Tau-GFP expressing cells (**Figure 4B-C**). Most strikingly, cells expressing P301L-Tau-GFP produced significantly more TIA1-positive stress granules per cell under basal conditions than WT-Tau-GFP expressing cells (p<0.05; **Figure 4B and 4D**). Together, these findings suggest that mutant tau enhances stress granule formation under basal and stressed conditions.

To examine endogenous *SNHG8* expression in the presence of mutant tau and with stress induction, *SNHG8* levels were monitored using RNAscope. Under basal conditions, *SNHG8* was significantly reduced in cells expressing P301L-Tau-GFP compared to WT-Tau-GFP (p<0.0001; **Figure 5**). With stress induction by culturing cells in Hank’s buffer, we observed a significant reduction of *SNHG8* in cells expressing WT-Tau-GFP or P301L-Tau-GFP compared to vehicle controls (p<0.0001; **Figure 5**). The most robust reduction in *SNHG8* was observed in P301L-Tau-GFP-expressing cells under stress conditions (**Figure 5**). Thus, stress induction and the presence of mutant tau are each sufficient to reduce *SNHG8* levels.

**Figure 5:**
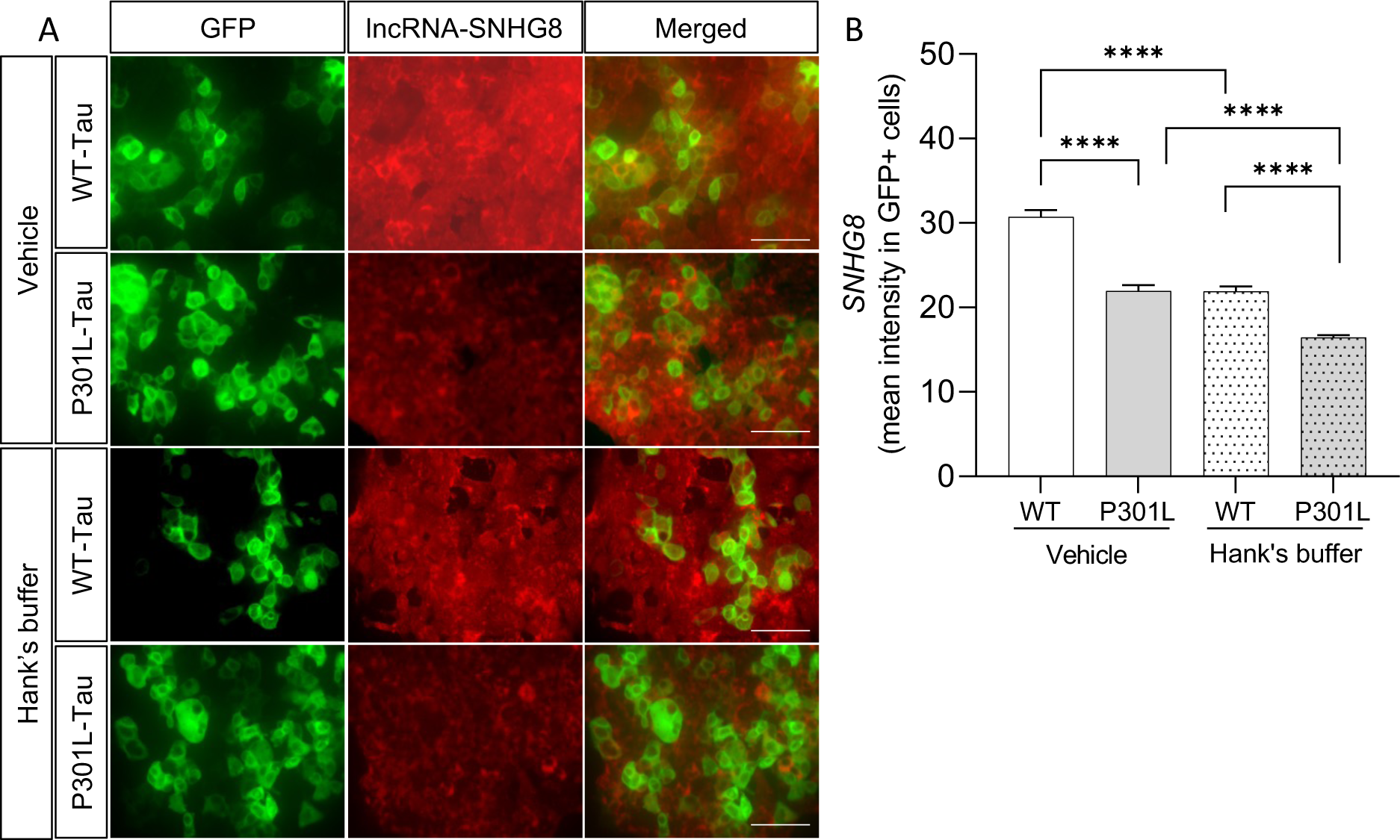
Mutant tau and stress lead to downregulation of *SNHG8*. A. RNAscope for *SNHG8* in WT-Tau-GFP and P301L-Tau-GFP-expressing cells under basal (vehicle) and stress (Hank’s buffer) conditions. GFP (green), *SNHG8* (red). White arrow, stress granules. Scale bar, 91um. B. Bar graph representing the quantification of mean intensity of *SNHG8* in GFP-positive cells. Data is representative of 4 independent experiments. Bar graphs represent mean ± SEM. Statistical significance was determined using a Student’s t-test. p***≤0.0001.

### SNHG8 blocks stress granule formation

We show that *SNHG8* interacts with tau and TIA1 and that mutant tau and stress induction regulate *SNHG8* levels. Thus, we asked if rescuing the expression of *SNHG8* could reduce stress granule assembly in P301L-Tau-expressing cells. HEK293-T cells were co-transfected with P301L-Tau and *SNHG8-*GFP or GFP vector control. This resulted in a 5-fold increase in *SNHG8* transcript levels (**Supplemental Figure 3**). Overexpression of *SNHG8* was sufficient to reduce the percentage of cells with TIA1-positive stress granules and the number of TIA-1 positive stress granules per cell (**Figure 6A-C**). The impact of *SNHG8* on stress granule formation may be driven by repression of TIA1 expression, as TIA1 protein levels are significantly reduced in P301L-Tau-expressing cells (**Figure 6A and 6D**).

**Figure 6:**
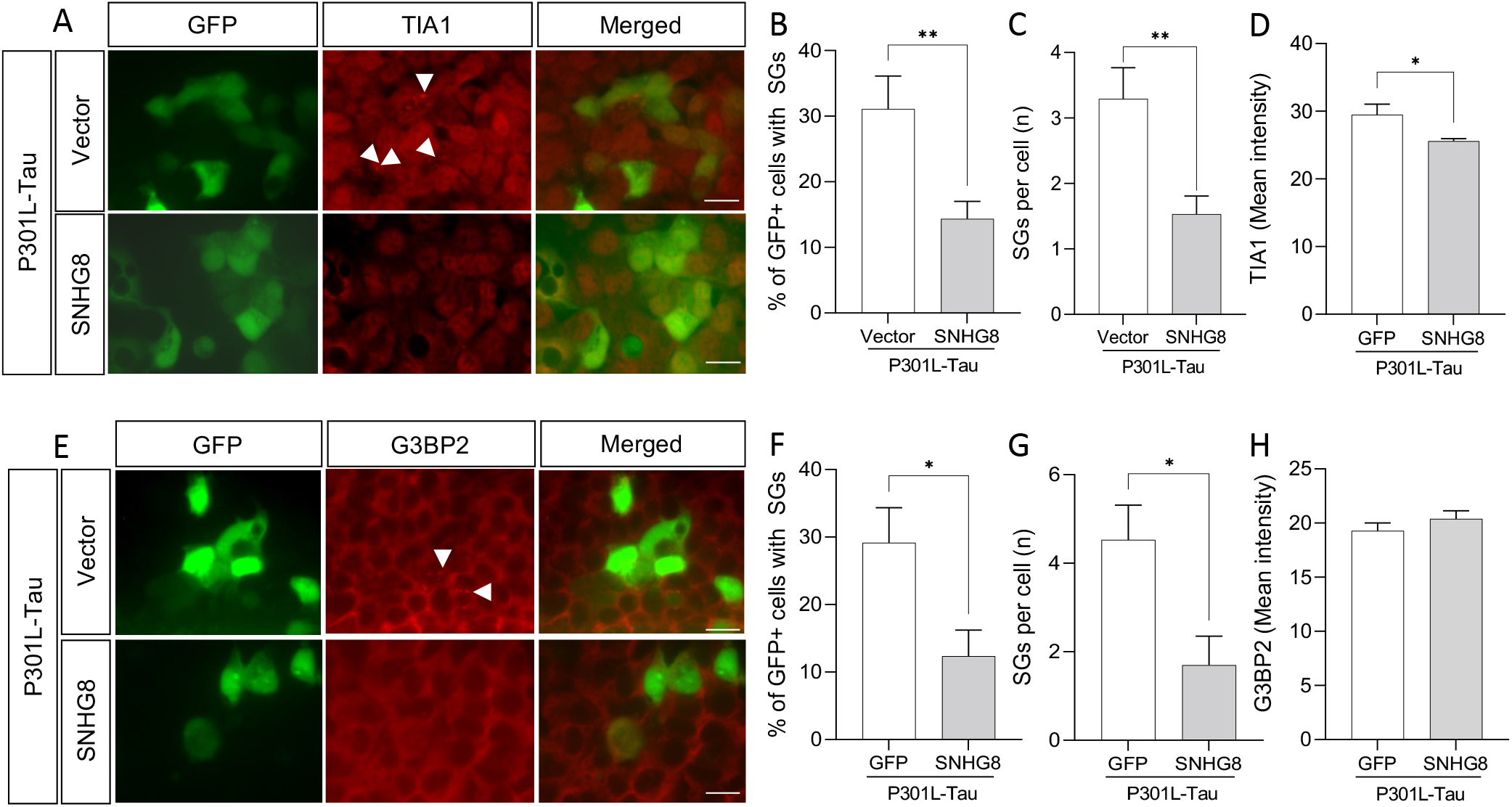
Overexpression of *SNHG8* inhibits stress granule formation in cells expressing mutant tau. Immunocytochemistry of HEK293-T cells co-overexpressing P301L-Tau with GFP or *SNHG8*-GFP under basal conditions. A. GFP (green), TIA1 (red). White arrow, stress granules. Scale bar, 50um. B. Graph representing the quantification of cells with TIA1-positive stress granules. C. Graph of the number of stress granules in GFP-positive cells. D. Graph representing TIA-1 levels. E. GFP (green), G3BP2 (red). White arrow, stress granules. Scale bar, 50um. B. Graph representing the quantification of cells with G3BP2-positive stress granules. C. Graph of the number of stress granules in GFP-positive cells. D. Graph representing G3BP2 levels. Data are representative of 4 independent experiments. Bar graphs represent mean ± SEM. Student’s t-test was performed to determine statistical significance. p*≤0.05, p***≤0.0001.

To determine whether *SNHG8* has a broad effect on stress granule formation, we evaluated G3BP2 levels, a stress granule assembly factor. *SNHG8* overexpression led to reduced G3BP2-positive stress granules in P301L-Tau overexpressing cells as compared to vector control (**Figure 6E-G**). However, *SNHG8* overexpression failed to change overall G3BP2 intensity (**Figure 6E-H**). Thus, *SNHG8* overexpression reduces stress granule formation without altering G3BP2 protein levels. Together, these studies suggest that *SNHG8* regulates stress granule formation by modifying TIA1 levels (**Figure 7**).

**Figure 7:**
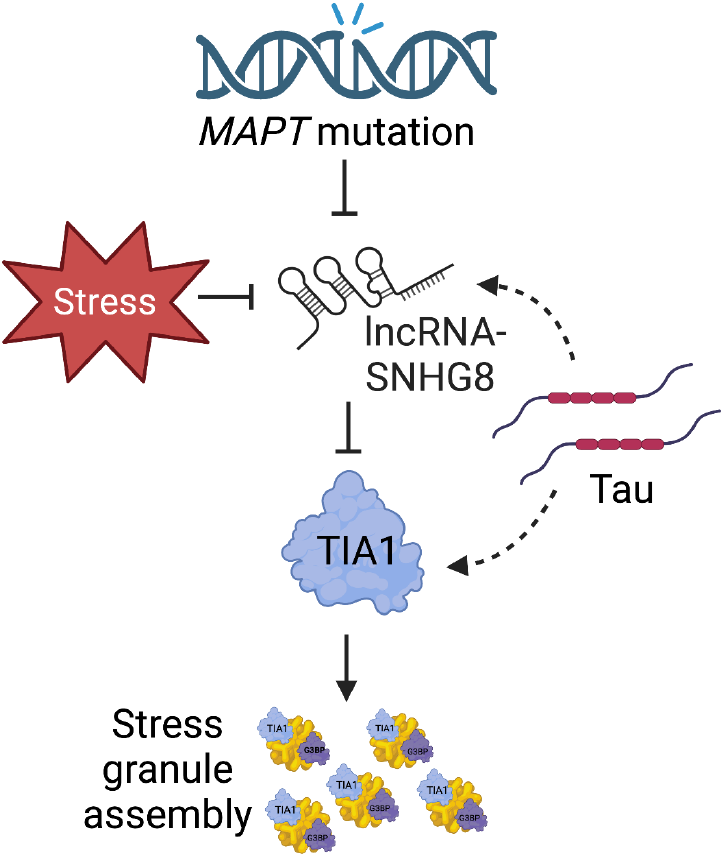
SNHG8 is a regulator of stress granule formation in tauopathy. Schematic of major findings. MAPT mutations or stress led to repression of SNHG8 expression. This repression limits SNHG8 interaction with Tau and enables Tau to interact with TIA-1 and form stress granules.

## Discussion

The goal of this study was to investigate the regulatory potential of lncRNAs in tauopathies. Patient-derived iPSCs have served as a powerful tool for studying the molecular and cellular mechanisms of neurodegeneration. Here, we identify 15 lncRNAs that are commonly differentially expressed across the *MAPT* mutations in human iPSC-derived neurons. These 15 lncRNAs function to regulate protein-coding gene expression in the human neurons and to interact with RNA-binding proteins involved in stress granule formation. Among these lncRNAs, *SNHG8* was significantly reduced in a mouse model of tauopathy and in the brains of patients with tauopathy, supporting a role for *SNHG8* in pathologic processes. We show that *SNHG8* interacts with tau, and overexpression of tau *in vitro* is sufficient to reduce *SNHG8* expression and induce TIA1-positive stress granule formation. Genetic manipulation of *SNHG8* leads to reduced stress granule formation suggesting that dysregulation of this non-coding RNA is a causal factor driving stress granule formation in tauopathies (**Figure 7**).

LncRNAs can alter gene expression by cis or trans mechanisms. We found that the commonly differentially expressed lncRNAs were highly correlated with coding genes commonly differentially expressed in the presence of *MAPT* mutations (Minaya et al. 2023). Gene enrichment analyses identified pathways related to Neurotrophin trk receptor signaling, Notch signaling, BDNF signaling, lipoprotein lipase activity, and axonal guidance. The 275 protein-coding genes are enriched in pathways associated with neuronal processes, synaptic function, and endolysosomal function (Minaya et al. 2023). However, here, we find that differentially expressed lncRNAs impact pathways that are restricted to those related to neuronal processes. Synaptic dysfunction has been widely reported among *MAPT* mutation carriers and in FTLD-tau (Nakamura et al. 2019; Tracy et al. 2022). Interestingly, the 15 lncRNAs are also associated with lipoprotein lipase activity, which point to a regulatory role in lipid metabolism. Cholesterol dyshomeostasis and the accumulation of lipid droplets have been reported in tauopathies (Glasauer et al. 2022). Thus, dysregulation of lncRNAs may contribute to several disparate phenotypes observed in tauopathies.

Dysregulation of RNA-binding proteins has been linked to various neurological disorders, including ALS, FTLD, AD, Huntington’s disease (HD), and Creutzfeldt-Jakob disease (CJD) (Gunawardana et al. 2015; B. Maziuk, Ballance, and Wolozin 2017). TARDBP gene encodes the TDP-43 protein, which is the primary component of pathological aggregates in most cases of ALS and 40% of cases of FTLD associated with progranulin haplo-insufficiency (Neumann et al. 2006). Similarly, mutations in FUS, hnRNPA1/B2, and other RNA-binding proteins have been associated with familial forms of motor neuron disorders (B. Maziuk, Ballance, and Wolozin 2017). Interaction of tau with RNA-binding proteins and ribosomes affects protein translation and RNA metabolism. In AD, the association of tau with RNA-binding proteins is increased, which may contribute to the dysregulation of RNA metabolism (Meier et al. 2016). LncRNAs have also been shown to interact with a wide range of RNA-binding proteins, which can impact posttranslational modification, stability, subcellular localization, and activity of interacting partners (McMillan et al. 2023; Yang, Wen, and Zhu 2015). The 15 common lncRNAs identified in our study were predicted to interact with several RNA-binding proteins that have been previously implicated in neurodegeneration. RNA-binding proteins including FUS, TARDBP, and TIA1 have been shown to contribute to pathology in FTLD-tau and FTLD-TDP (Ash et al. 2021; Gerstberger et al. 2014; Latimer et al. 2021; Montalbano et al. 2020; Urwin et al. 2010). Thus, studying the impact of lncRNAs across neurodegenerative disease may reveal novel disease mechanisms. Another hallmark of FTLD-tau is the presence of pathologic stress granules in neurons.

Stress granules are cytoplasmic complexes that form in response to nutritional stress, DNA damage, and proteostatic dysfunction (Kedersha and Anderson 2007; Mahboubi and Stochaj 2017; Vanderweyde et al. 2016). The liquid–liquid phase separation of stress granules is primarily driven by weak electrostatic, hydrophobic, and homo- and heterotypic protein–protein interactions between RNA-binding proteins that contain intrinsically disordered domains. Hence, identifying molecular drivers that affect the stability and assembly of stress granules is crucial to understanding basic molecular mechanisms of stress granule assembly and their role in the neuropathogenesis of FTLD-tau. The presence of stress granules has been reported in a mouse model of tauopathy and FTLD-tau patients (Vanderweyde et al. 2016). Under physiological conditions, tau has been shown to selectively co-partition with the RNA-binding protein TIA1, which consists of intrinsically disordered domains or prion-like domains to form aggregates (Ash et al. 2021; Wolozin and Ivanov 2019). However, molecular drivers involved in the regulation of stress granule assembly in tau neuropathology were unknown. Here, we provide evidence that *SNHG8* is a major regulator of TIA1-mediated stress granule formation. We find that when stress granules form, in the presence of mutant tau or in starvation conditions, *SNHG8* levels are reduced. Additionally, rescuing *SNHG8* levels reduces stress granule formation and TIA1 protein levels. These findings are consistent with prior observations that downregulation of TIA1 inhibits stress granule formation (B. F. Maziuk et al. 2018) and tau accumulation (Ash et al. 2021; Piatnitskaia et al. 2019; Vanderweyde et al. 2016). Interestingly, *SNHG8* was among the RNAs enriched in the interacting transcriptome of WT-Tau and P301L-Tau aggregates isolated from HEK293 biosensor cells (Lester et al. 2021). Together, we provide novel insights into the mechanisms of stress granule assembly in the neuropathology of FTLD-tau via the lncRNA *SNHG8*/TIA1 axis.

Small nucleolar RNA host genes (SNHGs) are a group of lncRNAs that contain introns and exons in their sequences and generate small nucleolar RNAs through alternative splicing. *SNHG8* is a newly identified type of small nucleolus host RNA belonging to the lincRNA family, located on chromosome 4q26 (Williams and Farzaneh 2012; Yuan, Yan, and Xue 2021). *SNHG8* is expressed by most cell types in the brain including neurons, oligodendrocytes, microglia, and astrocytes (Y. Zhang et al. 2014). *SNHG8* has been largely studied in the context of tumorigenesis, where it has been shown to promote the proliferation and invasion of cancer cells in gastric cancer, breast cancer, and ovarian cancer (Miao et al. 2020; Yu et al. 2021; Zou et al. 2021). The role of *SNHG8* in brain function and disease is not well investigated. Recently, *SNHG8* was reported to be involved in inflammation and microglial response by sponging miR-425-5p and SIRT1/NF-κB signaling (Tian et al. 2021) and in orthodontic tooth movement (OTM)(C. Wang et al. 2022). *SNHG8* has been associated with an inflammatory response: reducing *SNHG8*, which binds to HIF-1α, leads to free functional HIF-1α and activation of the downstream NF-κB pathway (C. Wang et al. 2022). The consideration of therapeutic strategies involving *SNHG8* replacement will require an evaluation of the landscape of *SNHG8* effects.

This study focused on those lncRNAs that were commonly differentially expressed across *MAPT* mutation types and associated with tau pathological events *in vivo*. This approach allowed us to focus on those lncRNAs that are most relevant to disease processes. However, lncRNAs are not fully annotated in many of the publicly available datasets, limiting our validation and translational potential. Therefore, additional investigation of lncRNAs in neurodegeneration will be important. Together, our study provides novel insights into the role of lncRNAs in pathological events leading to tauopathy. We show that lncRNA *SNHG8* is an interacting partner of tau, and *SNHG8* is a biological inhibitor of stress granule assembly via TIA1.

## Data Availability

All data produced in the present study are available upon reasonable request to the authors.

## Acknowledgements

We would like to thank the research subjects and their families who generously participated in this study. We thank Dr. Aimee Kao who generously provided the fibroblasts used to generate the GIH36C2 iPSC line (supported by the Rainwater Charitable Organization). This work was supported by access to equipment made possible by the Hope Center for Neurological Disorders, the Neurogenomics and Informatics Center, and the Departments of Neurology and Psychiatry at Washington University School of Medicine. Confocal images were generated on a Zeiss LSM 880 Airyscan Confocal Microscope, which was purchased with support from the Office of Research Infrastructure Programs (ORIP), a part of the NIH Office of the Director under grant OD021629. Funding provided by the National Institutes of Health (P30 AG066444, RF1 NS110890, U54 NS123985, K24 AG053435), Rainwater Charitable Organization (CMK), and UL1TR002345. The recruitment and clinical characterization of research participants at Washington University were supported by NIH P30AG066444 (JCM), P01AG03991 (JCM), and P01AG026276 (JCM). The UCSF Neurodegenerative Disease Brain Bank receives funding support from NIH grants P30AG062422, P01AG019724, U01AG057195, and U19AG063911, as well as the Rainwater Charitable Foundation and the Bluefield Project to Cure FTD. Diagrams were generated using BioRender.com.

