## Supplemental Figures for "Long non-coding RNA *SNHG8* drives stress granule formation in tauopathies"

### Supplemental Figure 1

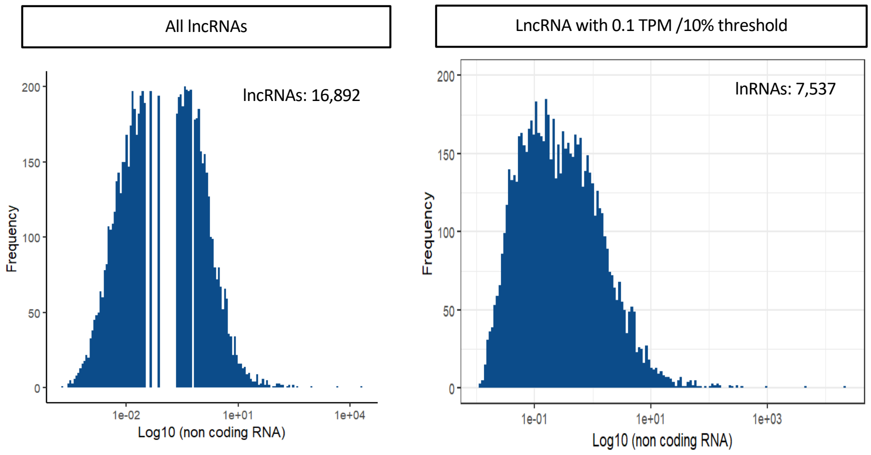


| Filter | Genes | Percentage |
|---|---|---|
| Total RNAs | 60,754 | 100% |
| Total number of non-coding RNAs | 16,892 | 26.8% |
| ≥0.1 TPM ≥10% threshold | 7,537 | 12.4% |

**Supplemental Figure 1: lncRNA distribution in sequenced neurons.** Left panel, distribution of lncRNA expression (including all lncRNAs). Right panel, after applying thresholds of 0.1 TPM expression in 10% of samples, histogram captures normal distribution. Table summarizing the numbers of genes at each stage. *TPM: transcripts per kilobase Million. Normalized for gene length and for sequence depth

### Supplemental Figure 2

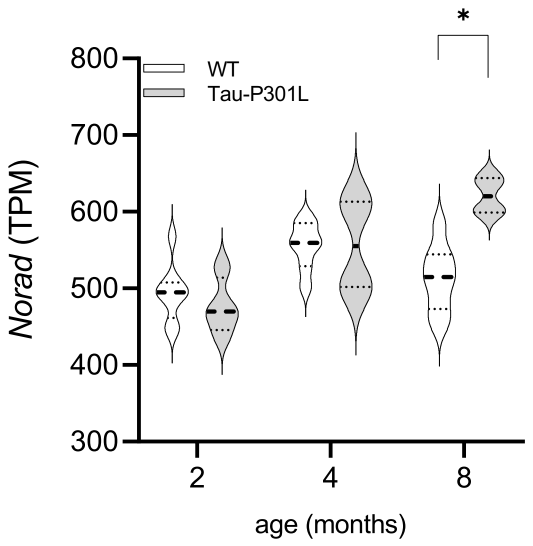


**Supplemental Figure 2: *Norad* is differentially expressed in *MAPT* p.P301L mice.** Normalized read counts (TPM) of *Norad* in WT mice and Tau-P301L mice. *, p<0.05.

### Supplemental Figure 3

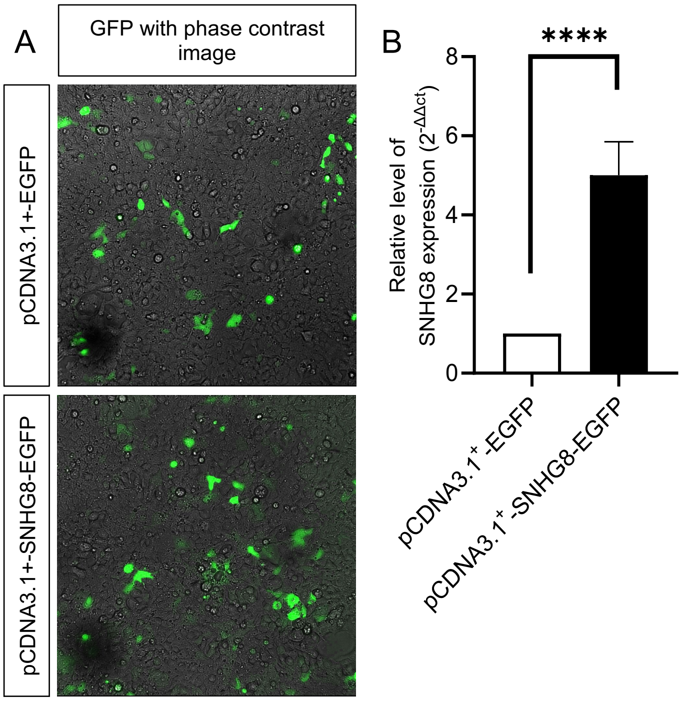


**Supplemental Figure 3: Ectopic expression of *lncRNA-SNHG8* in HEK293T cells**. A. Representative fluorescence images of HEK293T cells transfected with pCDNA3.1+-EGFP and pCDNA3.1+-SNHG8-EGFP plasmid constructs and images were taken 48hrs post transfection. The fluorescence images were overlayed with specific phase contrast image. B. Relative expression of *SNHG8* in HEK293T cells transfected with pCDNA3.1+-EGFP or pCDNA3.1+-SNHG8-EGFP plasmids. Expression normalized to GAPDH. n=4. Data are represented as mean ± SEM. **** $p < 0.0001$; unpaired Student's t-test.
